# Cost-effective sequence analysis of 113 genes in 1,192 probands with retinitis pigmentosa and Leber congenital amaurosis

**DOI:** 10.1101/2022.11.24.22282656

**Authors:** Daan M. Panneman, Rebekkah J. Hitti-Malin, Lara K. Holtes, Suzanne E. de Bruijn, Janine Reurink, Erica G.M. Boonen, Muhammad Imran Khan, Manir Ali, Sten Andréasson, Elfride De Baere, Sandro Banfi, Miriam Bauwens, Tamar Ben-Yosef, Béatrice Bocquet, Marieke De Bruyne, Berta de la Cerda, Frauke Coppieters, Pietro Farinelli, Thomas Guignard, Chris F. Inglehearn, Marianthi Karali, Ulrika Kjellström, Robert Koenekoop, Bart de Koning, Bart P. Leroy, Martin McKibbin, Isabelle Meunier, Konstantinos Nikopoulos, Koji M. Nishiguchi, James A. Poulter, Carlo Rivolta, Enrique Rodríguez de la Rúa, Patrick Saunders, Francesca Simonelli, Yasmin Tatour, Francesco Testa, Alberta A.H.J. Thiadens, Carmel Toomes, Anna M. Tracewska, Hoai Viet Tran, Hiroaki Ushida, Veronika Vaclavik, Virginie J.M. Verhoeven, Maartje van de Vorst, Christian Gilissen, Alexander Hoischen, Frans P.M. Cremers, Susanne Roosing

## Abstract

Retinitis pigmentosa (RP) and Leber congenital amaurosis (LCA) are two groups of inherited retinal diseases (IRDs) where the rod photoreceptors degenerate followed by the cone photoreceptors of the retina. A genetic diagnosis for IRDs is challenging since >280 genes are associated with these conditions. While whole exome sequencing (WES) is commonly used by diagnostic facilities, the costs and required infrastructure prevent its global applicability. Previous studies have shown the cost-effectiveness of sequence analysis using single molecule Molecular Inversion Probes (smMIPs) in a cohort of patients diagnosed with Stargardt disease and other maculopathies. Here, we introduce a smMIPs panel that targets the exons and splice sites of all currently known genes associated with RP and LCA, the entire *RPE65* gene, known causative deep-intronic variants leading to pseudo-exons, and part of the RP17 region associated with autosomal dominant RP, by using a total of 16,812 smMIPs. The RP-LCA smMIPs panel was used to screen 1,192 probands from an international cohort of predominantly RP and LCA cases. After genetic analysis, a diagnostic yield of 56% was obtained which is on par with results from WES analysis. The effectiveness and the reduced costs compared to WES renders the RP-LCA smMIPs panel a competitive approach to provide IRD patients with a genetic diagnosis, especially in countries with restricted access to genetic testing.

## Introduction

Inherited retinal diseases (IRDs) are a group of clinically and genetically heterogeneous disorders that are characterized by the dysfunction and subsequent death of the photoreceptor and/or retinal pigment epithelium (RPE) cells, which leads to reduced vision and can ultimately result in complete blindness. Phenotypic classification of IRDs is based on the affected cell type, the affected region within the retina, and the disease progression. For instance, retinitis pigmentosa (RP) is characterized by degeneration of rod photoreceptors and subsequently cone photoreceptors. This results in night blindness followed by peripheral vision loss and, for many persons, ultimately in blindness (Verbakel et al., 2018). In contrast, Leber congenital amaurosis (LCA) results in severe visual impairment or even complete blindness before the first year of life and is caused by degeneration of rod and cone photoreceptors, and often the RPE (den Hollander et al., 2008). The identification of genetic variants underlying IRDs is challenging as over 280 genes are currently known to be involved and pathogenic variants in many of these genes are known to cause multiple phenotypes (http://sph.uth.edu/retnet). Whole exome sequencing (WES) is often used in diagnostic facilities to identify causal variants explaining these phenotypes but the costs and the requirement for suitable infrastructure and bioinformatics support are still prohibiting its global applicability. An efficient and cost-effective targeted sequencing method is therefore desirable. Recently, single molecule Molecular Inversion Probes (smMIPs)-based targeted sequencing has been shown to fulfil this need in the field of IRDs. Using this approach, all genes associated with inherited macular degeneration (iMD) were sequenced, achieving a good overall detection and initial solve rate (Hitti-Malin et al., 2022).

In a similar approach, we have developed a smMIPs panel that targets all genes associated with the rod-dominated forms of the IRD spectrum. All genes known to be mutated in persons with RP and/or LCA, as well as genes associated with other rod-dominant IRDs such as congenital stationary night blindness (CSNB), gyrate atrophy, choroideremia, and Sørsby fundus dystrophy, were included. In contrast to standard WES without customized enrichment, the targeted smMIPs approach allows for enrichment and the sequencing of non-coding, deep-intronic regions of genes in which splice-altering variants have been detected previously. This was exemplified by an earlier smMIPs-based sequencing effort, in which the entire 128-kb *ABCA4* gene, including coding and non-coding regions, was sequenced in 1,054 Stargardt cases (Khan et al., 2020). In this study, a total of 13 novel causative deep-intronic variants (DIVs) were identified. Besides targeting DIVs, the smMIPs approach also allows for the investigation of other genomic regions of interest, such as the RP17 locus in which several different structural variants (SVs) have been identified causing autosomal dominant RP (RP ad) (de Bruijn et al., 2020). Finally, to identify cases eligible for available gene therapies, we have targeted the coding and non-coding regions of the *RPE65* gene.

Here, we present the outcome for 1,192 probands that underwent smMIPs-based sequencing using the RP-LCA smMIPs panel. We sequenced 360 probands in parallel per sequencing run in a highly cost-effective manner and obtained sequencing data that enabled the detection of single nucleotide variants (SNVs), small insertions and deletions (indels), and SVs, including copy number variants (CNVs), and revealed pathogenic variants underlying IRD in these individuals.

## Material and methods

### Gene selection and generation of the RP-LCA smMIPs panel

All genes implicated in RP and/or LCA, as well as genes associated with other rod-dominant IRDs (e.g. congenital stationary night blindness (CSNB)), were included in the design for smMIPs in order to completely target all rod-dominant IRDs (Supplemental Table 1). The selection of genes was based on the Retinal Information network online resource (https://sph.uth.edu/retnet/; accessed on 07-08-2020). Additionally, 417 smMIPs covering the RP17 locus, in which several pathogenic duplications and duplication-inversion events were identified, were added to the panel, and allowed detection of known and novel SVs at this locus (Supplemental Figure 1) (de Bruijn et al., 2020). *ZNF513* was included in the panel but was later withdrawn as a candidate gene and therefore not included in the final analysis.

For all genes included in the RP-LCA panel, the 5’ and 3’ untranslated regions (UTRs), exons, and alternative protein-coding exons were selected as targets. Additionally, all pseudo-exons (PEs) resulting from causal published DIVs, including 20 nucleotides (nt) upstream and downstream these PEs, as well as sequences resulting from exon skipping, intron retention, or with an effect on promoter activity, were included. A complete list of these targets and their genomic coordinates can be found in Supplemental Table 2. For *RPE65*, for which gene augmentation therapy is available (Russell et al., 2017), all intronic regions were also included as targets. Transcript numbers for the protein coding transcript (or the longest transcript) were selected from the Alamut Visual software version 2.13 (Interactive Biosoftware) and subsequently visualized using the UCSC Genome browser (Karolchik et al., 2004). All transcripts were evaluated for the presence of alternative protein coding exons and alternative 5’ UTRs using the Ensembl Genome Browser (GRCh37; Ensembl release 101) (Yates et al., 2020). Using the UCSC Genome Browser, hg19 (GRCh37) genomic coordinates were extracted and provided to Molecular Loop Biosciences, USA (Haeussler et al., 2019). A total of 16,812 smMIPs were designed to cover the regions described above with flanking regions of at least 20 nt on both the 5’ and 3’ ends of the provided regions, resulting in a total of 453,462 nt that are covered by the RP-LCA smMIPs panel.

### smMIPs design

The Molecular Loop Biosciences’ smMIPs design includes 225 nt captured regions that are flanked by a 20 nt extension and a 20 nt ligation probe arm at the 5’ and 3’ end, respectively. All smMIPs are dual-indexed using two 10-nt unique index primer sequences, or ‘barcodes’, that act as a patient barcoding system to generate uniquely tagged libraries. To tag each individual smMIP, two 5 nt Unique Molecular Identifiers are included next to the probe arms and are used to detect duplicate reads and enable the detection of unique reads.

### Sample selection and preparation

Prior to shipment to the host institution, collaborators prepared the DNA samples. First, DNA concentrations of the samples were quantified using the Qubit dsDNA HS assay kit (Thermo Fisher, US), according to manufacturer’s instructions. Second, DNA was diluted to a concentration of 16.7 ng/μl and 100 ng of DNA was loaded on a 0.8% agarose gel flanked by uncut lambda DNA (25, 50, 100, 200, and 400 ng) and 0.5 μg of a 1-kb ladder. Based on this gel, the DNA concentration was compared to the uncut lambda DNA and, subsequently, labeled as high or low molecular weight (MW) DNA based on the size distribution. The DNA was considered high MW when the DNA fragment was ≥ 23 kilobases (kb) and low when ≤ 23 kb and appeared as a smear on the gel. DNA samples of high MW were plated into a 96-well capture plate and subsequently pre-treated by incubating the DNA at 92°C for five minutes to briefly shear the DNA prior to library preparation. Next, DNA samples of low MW were added to the capture plate. Additionally, each plate contained six positive controls (five on every fourth plate) and one non-template control (NTC) containing only water.

### Library preparation

Sequencing libraries were generated as described previously (Hitti-Malin et al., 2022). The High Input DNA Capture Kit, Chemistry 2.3.0H (Molecular Loop Biosciences, Inc), was used according to Protocol version 2.4.1H. In short, smMIPs were hybridized for 18 hours followed by the fill-in reaction to circularize the probe and, subsequently, a combined clean-up and PCR step. Prior to pooling of all samples, the appropriate size (413 bp) of each individual product was evaluated by agarose gel electrophoresis. The pooled library was purified using bead purification and quantified using the Qubit Fluorimeter and the TapeStation system to assess library concentration and fragment sizes, respectively.

### Sequencing

Four sequencing library pools, generated with the High Input DNA capture kit, were combined in an equimolar fashion to form one 100 μl mega-pool of 1.5 nanomole (nM). This pool was denatured according to Illumina’s NovaSeq 6000 System Denature and Dilute Libraries Guide, yielding a 300 pM library. Each library was sequenced by paired-end sequencing on the NovaSeq 6000 platform (Illumina, California, USA) using SP reagent kits v1.5 (300 cycles).

### Variant calling and annotation

All reads generated by the NovaSeq 6000 run were converted into raw sequencing data files (FASTQ) using bcl2fastq (v2.20). These files were subsequently processed using a bioinformatics pipeline developed in-house, as described previously (Khan et al., 2019). In short, the random identifiers were removed from the sequencing reads and added to the read identifier for later use. After exclusion of duplicate reads, the remaining reads were added to patient specific BAM files based on the index barcoding system. In order to determine the overall average smMIPs coverage, forward and reverse read were combined and subsequently divided by two.

### Average coverage per nucleotide

To determine the number of reads covering each nucleotide in sequencing run 01, the base calls of aligned reads to a reference sequence were counted in BAM files corresponding to individual probands using the ‘pileups’ function of SAMtools (Li et al., 2009). Data was obtained using the following parameters: minimum mapping quality = 0, minimum base quality = 12, anomalous read pairs were discarded, overlapping base pairs from a single paired read as a depth of 1 were counted. An average coverage per nucleotide was generated for each nucleotide position across all samples sequenced in RP-LCA run 01, followed by an average coverage for all genes/loci targeted in the RP-LCA panel. The average coverage per nucleotide for *RPGR* was calculated excluding exon 15 of the *RPGR-ORF15* transcript, and the coverage of exon 15 of the *RPGR-ORF15* transcript was determined independently. Coverage plots for all reads across each gene/locus were generated. The average coverage per nucleotide was used to assess whether regions were poorly covered (≤10 reads), moderately covered (11-49 reads), or well-covered (≥50 reads).

### Variant prioritization and classification

CNV analysis was performed for all samples using an Excel script described previously (Khan et al., 2020). We presumed a deletion when six (or more) consecutive smMIPs with a normalized coverage across all samples in that run was equal to or smaller than 0.65. Conversely, a duplication was assumed if six (or more) consecutive smMIPs yielded a normalized coverage of ≥ 1.20.

Subsequently, all SNVs and indels were evaluated. First, previously published pathogenic DIVs were included in the filtering and prioritization steps. Secondly, all homozygous and heterozygous variants with an individual minor allele frequency (MAF) of ≤ 0.5% in genes associated with autosomal recessive IRDs and all heterozygous variants in genes associated with autosomal dominant IRDs with a MAF of ≤ 0.1% were assessed. MAFs were obtained from the Genome Aggregation Database (gnomAD v2.1.1; 125,748 exomes and 15,708 genomes), as well as from an in-house WES cohort consisting of data from 24,488 individuals with a large variety of clinical phenotypes. All three MAFs needed to meet the cut-offs described above. Variants that were called in ≥ 80% of the sequencing reads were considered homozygous and variants that were called in 35-80% of the sequencing reads were considered heterozygous. Variants that were called in ≥ 10% of probands included in a sequencing run (i.e. ≥ 38 probands) and were detected with less than 10 reads across that genomic position were excluded from further analysis.

Prioritization of variants was based on variant types, predicted protein effect, and pathogenicity scores. First, all stop gain, stop loss, frameshift, start loss, and canonical splice site variants were considered. Thereafter, in-frame insertions and/or deletions were assessed followed by missense variants that met the pre-defined thresholds of all three *in silico* pathogenicity prediction tools. Namely, PhyloP (threshold: ≥2.7, range: −14.1 – 6.4), CADD-PHRED (threshold: ≥15, range 1 – 99), and Grantham (threshold: ≥80, range 0 – 215) (Grantham, 1974; Pollard et al., 2010; Kircher et al., 2014).

Missense variants that met either one or two thresholds were prioritized thereafter. Subsequently, all variants were investigated using SpliceAI, except for variants affecting the canonical splice acceptor (+1 and +2 position) and donor splice sites (−1 and −2 position) (Jaganathan et al., 2019). Variants with a predicted delta score ≥ 0.2 (using the default settings with a window of −50 bp to +50 bp) on any of the four parameters (acceptor gain, acceptor loss, donor gain, or donor loss) were prioritized. All non-canonical splice-site (NCSS), near-exon variants and DIVs were investigated using *in silico* tools available via Alamut Visual. Splice Site Finder-like (Zhang, 1998), MaxEntScan (Yeo and Burge, 2004), NNSPLICE (Reese et al., 1997), and GeneSplicer (Pertea et al., 2001) were utilized to predict the effect on splicing according to parameters described before (Fadaie et al., 2019). ESE finder was used to predict the effect on exon splicing enhancers (Cartegni et al., 2003).

Using the ACMG/AMP classification system, all variants were assigned one of five classes: class 1 (benign), class 2 (likely benign), class 3 (variant of uncertain significance, or VUS), class 4 (likely pathogenic) or class 5 (pathogenic) (Richards et al., 2015). These classes were assigned according to the ACMG-AMP guidelines using the Franklin Genoox Platform (https://franklin.genoox.com/, accessed before November 2022). For *ABCA4*, the severity scores as published in Cornelis *et al*., were used to reach a final classification instead of the ACMG classification (Cornelis et al., 2022). For RP, we considered the proband to be very likely solved by a combination of moderate and/or severe *ABCA4* alleles. Mild *ABCA4* alleles, although sometimes classified as either class 3, 4, or 5, were deemed not to be causative for RP in this study.

Each proband was assigned an outcome indicating whether the proband was genetically “very likely solved”, “possibly solved”, or “unsolved”. Segregation analysis was not performed in this study and, therefore, no definitive “solved” label could be assigned. When at least two variants in a given gene were observed, they were listed in two alleles, although segregation analysis was not performed. Compound heterozygosity therefore was not proven and also was not added as proof for the ACMG classification. All modes of inheritance were taken into consideration when assessing the prioritized variants identified in a proband. When a class 4 or 5 variant was detected in a homozygous state in genes known to be associated with autosomal recessive IRDs, the proband was considered “very likely solved”. The genomic region in which the homozygous variant was detected was subsequently assessed for potential heterozygous deletions. In cases with two heterozygous variants in a gene associated with an autosomal recessive retinal disease, a combination of class 4 and/or 5 variants were sufficient to assign a “very likely solved” verdict. This was also the case for probands in which one class 5 and one class 3 variant was identified. Probands with one class 4 and one class 3 variant or two compound heterozygous class 3 variants were deemed “possibly solved”. Probands with a heterozygous class 4 or 5 variants observed in a gene associated with autosomal dominant inheritance were assumed to be “very likely solved”, whereas a proband with a single heterozygous class 3 variant remained genetically “unsolved”.

### Minigene analysis

Minigene analysis was performed as previously described (Sangermano et al., 2018; Verbakel et al., 2019). In short, the regions of interest of the genomic DNA sample was amplified by primers that contain attB1 and attB2 tags at their 5⍰ end to facilitate Gateway cloning. After obtaining the entry clone, the wild-type and mutant construct containing the *RPE65*:c.675C>A variant were separately inserted into the pCI-NEO-RHO Gateway-adapted vector to generate wild-type and mutant minigenes. Both minigenes were independently transfected into HEK293T cells and after 48⍰h of incubation, mRNA was isolated and amplified by RT-PCR with primers in the flanking RHO exon 3 and 5 regions. All primers used for this splice assay are available upon request. Fragment sizes were assessed using gel electrophoresis and identified using Sanger sequencing.

### Ethical considerations

The study adhered to the tenets of the Declaration of Helsinki and was approved by the local ethics committee of the Radboud University Medical Center (Nijmegen, The Netherlands). Written informed consent was obtained from patients prior to DNA analysis and inclusion in this study.

## Results

Prior to sequencing 360 probands in a single sequencing run, a test run was performed including 32 control samples, harboring a total of 22 CNVs and 19 SNVs, together with 15 genetically unsolved probands. This test run was used to assess the average coverage across all targets and the performance of the RP-LCA smMIPs pool. A total of 496,610,877 reads were obtained after exclusion of duplicate reads, averaging at 10,566,189 reads per proband. An average of 629 reads per smMIP was obtained across the entire pool of 16,812 smMIPs and, since all nucleotides are covered by eight smMIPs on average, each nucleotide was covered by approximately 5,032 smMIPs on average. As this confirmed solid read characteristics, we continued to increase the number of probands sequenced in a single run to 360 cases whilst maintaining adequate read numbers to call variants (approximately 629 reads per nucleotide on average). Since even read coverage was achieved, no rebalancing of the smMIPs pool was required (Figure 1). All previously identified SNVs and CNVs from the positive controls could be detected and prioritized correctly, validating our filtering and prioritization procedure.

**Figure 1:**
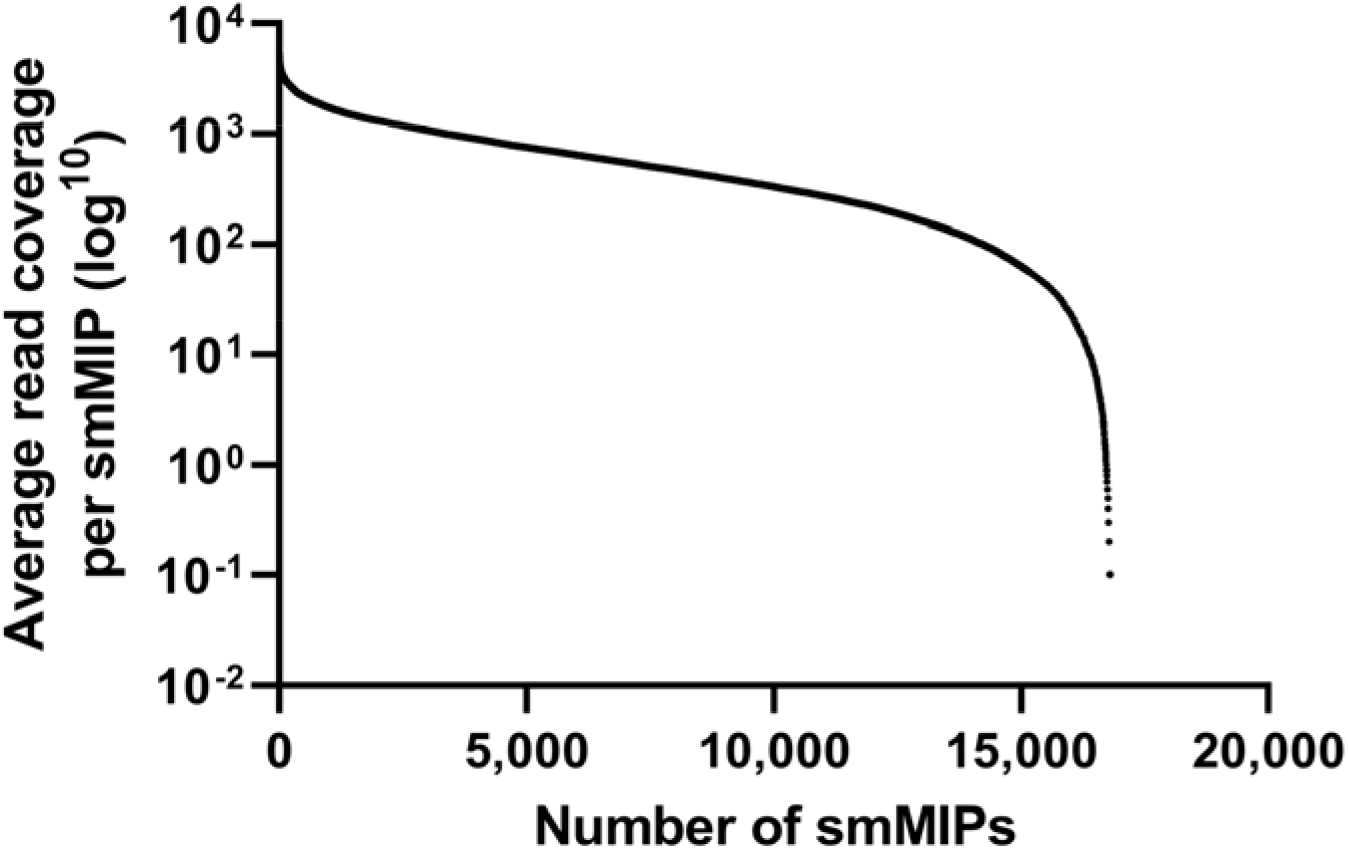
Average read coverage per smMIP in the RP-LCA panel.

In the first complete run (RP-LCA run 01), a total of 360 unsolved cases and 20 controls containing known CNVs, were sequenced. A total number of 596,889,627 reads were obtained after exclusion of duplicate reads. The amount of reads per proband was 1,570,762 on average and an average of 93 reads per smMIP was obtained. Using pileups data, we extracted the amount of reads covering each individual nucleotide and observed an average coverage per nucleotide of 374x. Nucleotides that were covered by ≥ 50 reads were considered well covered, nucleotides covered by 11 to 49 reads moderately covered, and nucleotides covered by ≤ 10 reads were deemed to be poorly covered. In RP-LCA run 01, 431,878 nt were well covered (95.8%), 14,932 nt (3.3%) were moderately covered, and 3,832 nt (0.9%) were poorly covered. From this, we calculated the coverage per target gene (Figure 2). We observed that the last exon of the *RPGR-ORF15* transcript, together with the *PRCD, NYX, SAMD11*, and *WDR34* genes, were the five genes/regions with the lowest average nucleotide coverage (72x, 152x, 156x, 157x, and 174x, respectively). The coverage of these genes/regions exceeded our threshold to be considered sufficient for variant calling (i.e. 50x). However, the validity of variants called in the *RPGR-ORF15* transcript was difficult to assess because of the repetitive nature of the region and would therefore need additional long-read sequencing validation.

**Figure 2:**
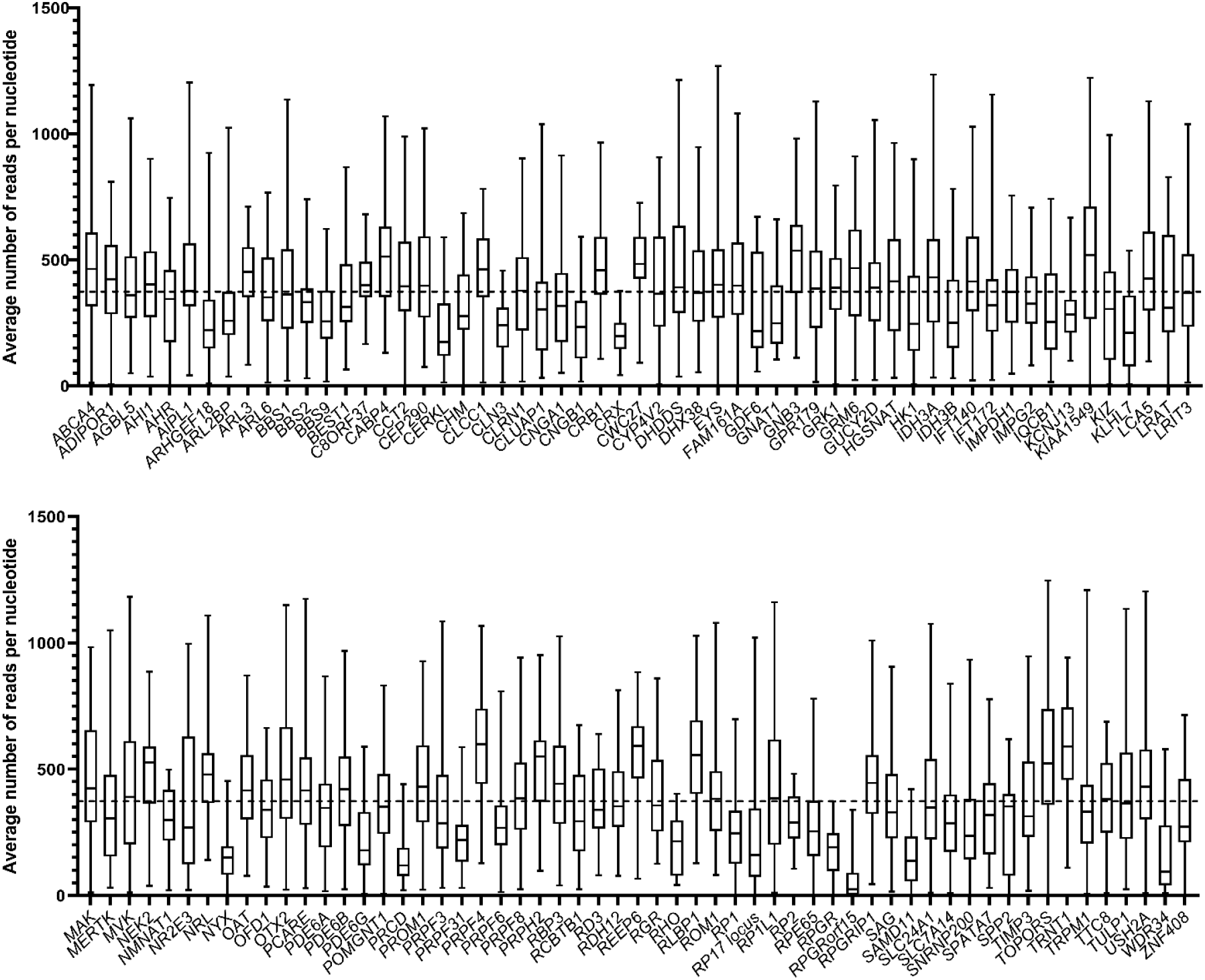
Average coverage per nucleotide. The average number of reads per nucleotide for each individual gene are indicated by the black horizontal line within the box, the range is indicated by the whiskers. The horizontal dashed line depicts the average overall coverage (374x).

To further validate our CNV analysis and SNV prioritization and classification workflow, we included another group of 63 probands diagnosed with either RP or LCA and previously screened by a MIPs panel targeting 108 genes associated with IRD (Weisschuh et al., 2018; Sharon et al., 2020). Using our workflow, we obtained a very likely or possibly solved verdict for 40 probands (Supplemental table 3). Of those 40, the variants detected in 28 probands were in concurrence with previous findings (group 1), nine were solved by variants previously not detected (group 2), three probands that were considered to be possibly solved through our workflow, were solved by variants in other genes (group 3). Of those three genes, two genes were not covered by our panel, and one variant in *PRPF8* (c.3394_3396delAAG, p.(Lys1132del)) was detected but excluded based on our workflow since we did not consider class 3 variants in autosomal dominant genes. Eight out of 22 probands that were considered unsolved after our analysis, were known to be genetically solved (group 4). Of those eight, five probands were solved with variants in genes not included in this panel since they are associated with other IRD phenotypes. Furthermore, one duplication was not detected in our CNV analysis, one SNV was detected but only with one read, and therefore excluded, and one proband was considered solved by a class 3 variant in an AD gene.

To enable a focused analysis, we took forward probands that were submitted with an RP (83.7%), LCA (8.7%) or retinal dystrophy (RD, 7.6%) phenotype. After exclusion of 32 samples that failed library preparation, a total of 1,191 probands were analyzed from five sequencing runs. After CNV and SNV analysis, 566 probands were considered very likely solved (47.5%) and 100 probands were considered possibly solved (8.4%), respectively. This resulted in a diagnostic yield of 55.9% when combining these groups (Supplemental Table 4). The 666 very likely and possibly solved probands could be explained by variants in 76 genes (Figure 3). For probands submitted with an RP phenotype (n=573), the most prevalent mutated genes were *USH2A* (17.3%), *EYS* (9.6%), *RHO* (5.1%), *RP1* (4.7%), and *PDE6A* (4.2%). Probands with an LCA phenotype (n=57) were mostly genetically explained by pathogenic variants residing in *CRB1* (14.5%), *ABCA4* (12.7%), *RPGRIP1* (9.1%), *CEP290* (5.5%), *GUCY2D* (5.5%), *RDH12* (5.5%) and *TULP1* (5.5%) while those with an RD phenotype (n=36) could be genetically explained by variants in 10 different genes (each in two cases) *ABCA4, CRB1, EYS, GUCY2D, NRE2E3, PROM1, PRPH2, RLPBP1, TRPM1*, and *USH2A* (all 3.6%).

**Figure 3:**
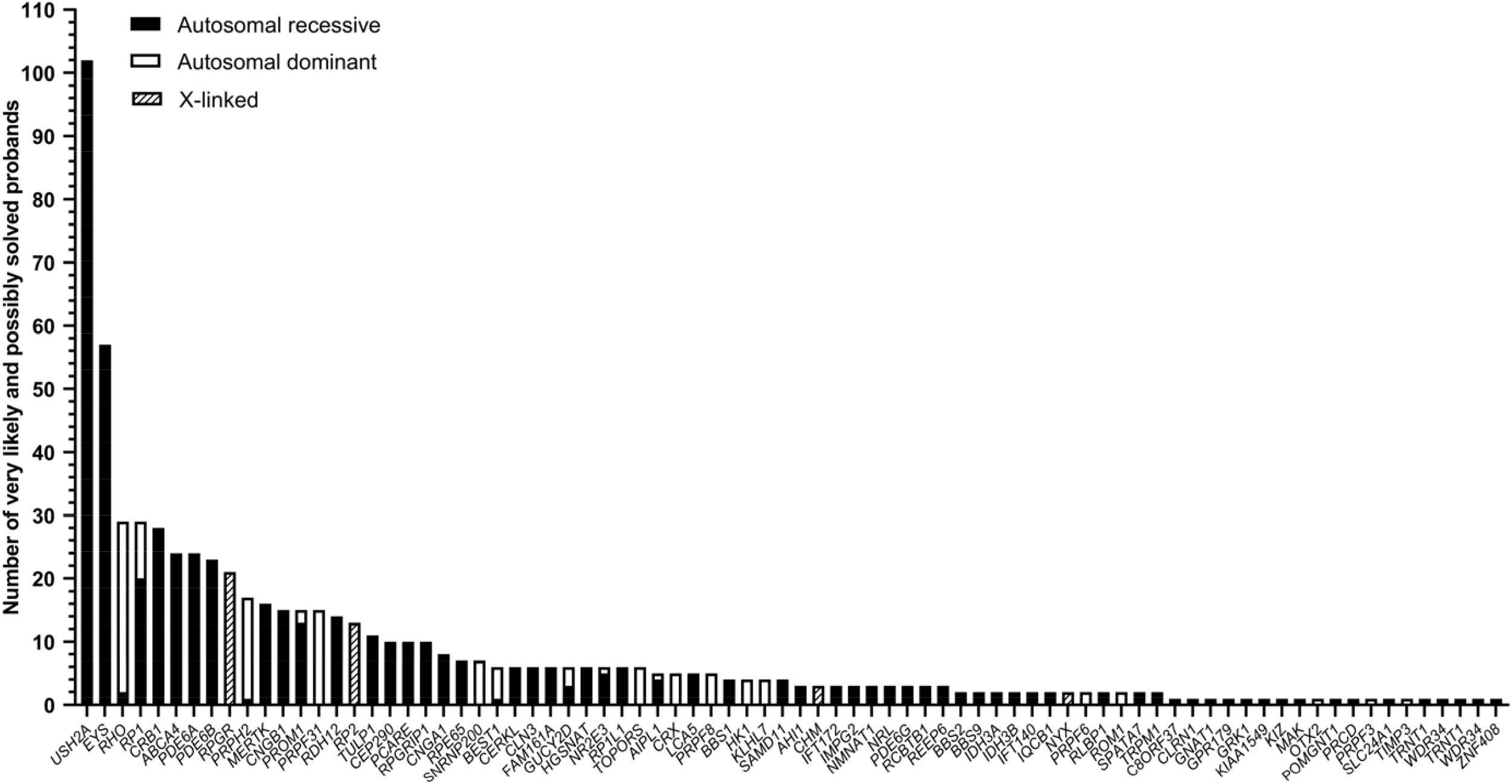
Number of solved cases per individual gene. For all genes, the inheritance mode is depicted in either green (autosomal recessive), blue (autosomal dominant), or orange (X-linked inheritance).

For CNV, SNV, and indel analysis, all possible phenotypes and modes of inheritance were considered, regardless of the details provided by the collaborator. In total, 40 CNVs and one SV were identified in 19 genes amongst all cases. Twenty-five heterozygous deletions, nine homozygous deletions, three hemizygous deletions, two heterozygous duplications, and one SV in the RP17 locus were detected by CNV analysis (Supplemental Table 5). All probands with heterozygous CNVs could be genetically explained by an additional pathogenic SNV on the presumed second allele. All class 3, 4, and 5 SNVs, CNVs, and indels were taken forward if detected in a homozygous or compound heterozygous state in a gene associated with autosomal recessive inheritance. In this way, 509 probands were possibly (85 probands) or very likely (423 probands) solved by assumed compound heterozygous (300 probands) or homozygous variants (210 probands) (Supplemental Table 6). For variants in genes associated with autosomal dominant inheritance, only class 4 and 5 SNVs and indels were assessed. All variants that were very likely (108 probands) or possibly solving (9 probands) a case were listed in Supplemental Table 7. Moreover, all class 3, 4, and 5 variants identified in X-chromosomal genes were also analyzed. All variants that were considered to possibly (6 probands) or very likely (33 probands) solve the proband are listed in Supplemental Table 8. In this study, 275 variants were detected that were not previously reported in literature. All variants and cases have been uploaded into the respective Leiden Open Variation Databases.

Among the called SNVs, we were able to detect known splice-site altering variants published previously in literature as they were included as targets for the RP-LCA smMIPs panel. We identified five probands with the deep-intronic c.2991+1655A>G variant in *CEP290* (den Hollander et al., 2006), one proband with the c.1374+654C>G in *PRPF31* (Frio et al., 2009), and one proband with the c.7595-2144A>G variant in *USH2A* (Vache et al., 2012). The probands in which the *CEP290* c.2991+1655A>G and *USH2A* c.7595-2144A>G variants were detected, could be genetically explained as they also carried a (likely) pathogenic variant on the assumed second allele. In proband 067528, one of the two probands carrying the c.1374+654C>G variant in *PRPF31*, a homozygous deletion of exon 4 in *BBS9* was also identified, ultimately resulting in a frameshift (p.(Gly89Asnfs*4)). For this individual, the *PRPF31* variant was listed as a secondary finding but could not be excluded for having an additional effect on the probands phenotype. All three DIVs were shown to lead to pseudo-exon inclusion and have been published previously (den Hollander et al., 2006; Frio et al., 2009; Vache et al., 2012; Fadaie et al., 2021).

For proband DNA13-01427, we detected the pathogenic c.886dup (p.(Arg296Lysfs*7) and the synonymous c.675C>A variant that was classified as VUS in *RPE65*. SpliceAI predictions for c.675C>A yielded a delta score of 0.39 and a donor loss with a delta score of 0.31 suggesting putative skipping of exon 7. Because of the therapeutic relevance of *RPE65*, the variant was assessed in a minigene splice assay with a genomic DNA insert of 3.8 kb containing *RPE65* exons 6 through 10 (figure 4A). The wild-type construct showed the expected wild-type fragment of 907 nt and also showed a fragment of 825 nt corresponding to the skipping of exon 7 (82 nt)suggesting natural exon 7 skipping (figure 4B). A similar phenomenon was observed in control photoreceptor precursor cells (PPCs) and pure cultures of control retinal pigment epithelium (RPE) cells (data not shown). RT-PCR analysis of the RNA resulting from the *RPE65* c.675C>A minigene, showed the same fragment of 825 nt corresponding to the skipping of exon 7 which leads to frameshift variant after 4 amino acids (p.(Asp215Valfs*4)). The RNA products resulting from the transfection of the mutant *RPE65* c.675C>A showed with no remaining wild-type RNA and therefore the allele was classified as severe (figure 4B). Adding this functional evidence to the ACMG classification resulted in a re-classification as pathogenic. Segregation for both variants was confirmed in both parents for this proband. In total from 1,192 probands, seven probands were very likely or possibly solved by pathogenic variants in *RPE65*.

**Figure 4:**
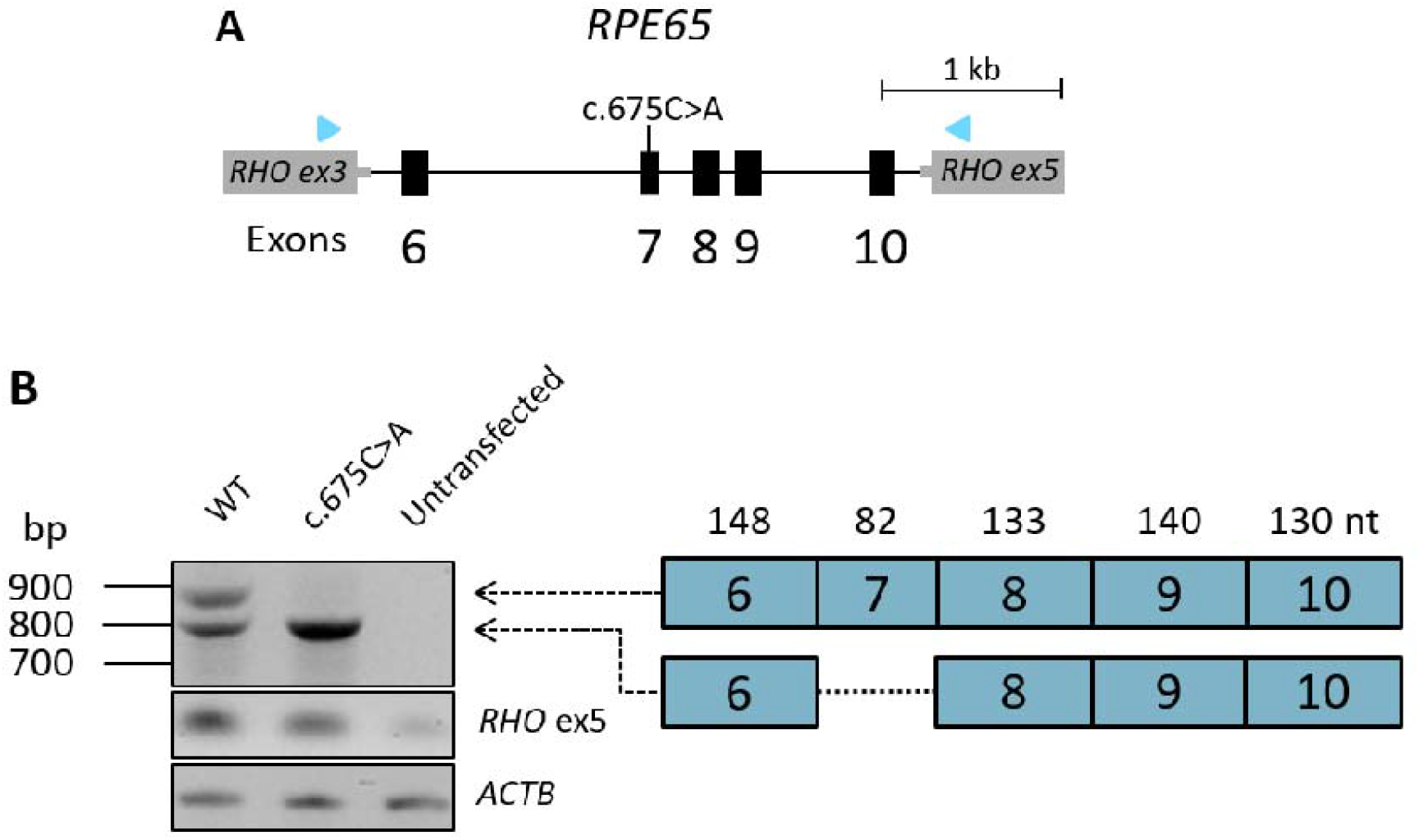
Minigene analysis of the splicing effect of *RPE65:*c.675C>A. A) A minigene construct containing either wild-type sequence or the mutant c.675C>A variant in exon 7 was generated spanning exons 6-10 of *RPE65* and was flanked by exons 3 and 5 of the *RHO* gene in the pCI-NEO-RHO vectors B) After gel electrophoresis of the RT-PCR product, we observed two fragments in the wild-type situation. One fragment of 907 nt and one fragment of 825 nt corresponding to the skipping of the 82 nt exon 7. For 675C>A, we exclusively observed the 825 nt fragment, suggesting complete skipping of exon 7.

In addition to known DIVs, we also targeted the RP17 locus in the RP-LCA panel. SVs in this locus have been shown to cause RP ad (de Bruijn et al., 2020). We identified two probands with duplicated regions of the RP17 locus. In proband 067984, we could confirm that the SV was identical to the UK-SV2 variants published previously (de Bruijn et al., 2020). The second proband (RP ar phenotype) did not harbor a known SV from the de Bruijn *et al*., study [5]. Using SNP array analysis, it was revealed that this SV it unlikely to cause alteration of the topologically associating domain (TAD) organization of the RP17 locus and is therefore, in combination with the phenotype, considered to be benign (data not shown).

## Discussion

Using the RP-LCA smMIPs panel, we sequenced all exonic regions of the genes associated with RP and LCA and, additionally, the intronic regions of the *RPE65*, all previously published DIVs in RP/LCA genes, as well as the RP17 locus associated with RP ad in 1,192 probands. After CNV analysis and prioritization of SNVs and indels, we very likely or possibly solved 666 probands (55.9%) with causative variants distributed over 76 genes. When comparing the diagnostic yield of groups of probands originating from different collaborators, a range of 51%-63% was observed. A previous study that used WES to analyze 266 probands with various types of IRDs, had reached a slightly higher diagnostic yield of 63% in a RP subgroup (Haer-Wigman et al., 2017). This difference can possibly be attributed to differences in the genetic screening methods that probands had received prior to inclusion in our RP-LCA smMIPs sequencing. For example, in a group of probands originating from Italy, which had previously undergone APEX microarray analysis, we obtained a diagnostic yield of 51% (26/51 cases) after our analysis, whereas another cohort from France, that received no prior genetic screening showed a diagnostic yield of 63% (116/184 cases).

In the RP subgroup, the top five genes in which variants that solved the proband were detected consisted of *USH2A, EYS, RHO, RP1*, and *PDE6A*. This set of genes is comparable to the genes listed in a recent review by Verbakel *et al*. on RP, which includes a shortlist of frequently mutated genes (*USH2A, RPGR, EYS, RHO*, and *RP1*) (Verbakel et al., 2018). The large proportion (approximately 4x higher than expected) of probands solved by variants in *ABCA4* is likely explained by the extensive efforts on determining the complex pathogenicity of variants in *ABCA4* compared to other genes (Al-Khuzaei et al., 2021; Cornelis et al., 2022; Lee et al., 2022). The underrepresentation of variants detected in genes on the X chromosome might be explained by more extensive targeted testing of X-linked IRD genes such as *RPGR* (prior to smMIPs sequencing) in probands with X-linked diseases. As previously mentioned, variants in *USH2A* solve the largest number of cases in this study. Common variants such as c.2276G>T and c.2299del were observed in 28 and 17 cases, respectively. Furthermore, we could possibly or very likely solve 13 probands with variants in very rarely mutated disease genes such as *IDH3A* (2x), *IDH3B* (2x), *KIAA1549* (1x), *KIZ* (1x), *RCBTB1* (3x), *REEP6* (3x), and *TRNT1* (1x). For instance, *KIAA1549* was determined to be causative in only three probands from three families previously (Abu-Safieh et al., 2013; de Bruijn et al., 2018). Here, we detected a novel homozygous c.4427dup variant (p.(Glu1477Glyfs*3)), which very likely solves proband 067904.

Variant prioritization was completed without taking into consideration the phenotype and possible mode of inheritance provided by the collaborator. This genotype-first approach implies that the variant prioritization may establish an unexpected genetic outcome for a proband, which may not fit the phenotype and/or inheritance pattern provided by the collaborator. For instance, in seven probands diagnosed with LCA we observed pathogenic variants in *ABCA4*. These probands were very likely solved based on our workflow, but variants in *ABCA4* have never been associated with LCA previously. Six out of seven probands carried two protein-truncating variants and one proband carried a homozygous missense variant (p.(Glu1022Lys)), which was deemed to have a moderate/severe effect on ABCA4 function (Cornelis et al., 2022). The phenotype of the proband in which the latter variant was found was re-assessed and Stargardt disease was considered to be more likely. Segregation analysis and in-depth clinical investigation could confirm this new genotype-phenotype correlation. Furthermore, genes associated with autosomal dominant inheritance, such as *PRPF31*, can show reduced penetrance and therefore a proband with a proposed autosomal recessive disorder might be explained with a variant in this gene (McGee et al., 1997). In fact, of the 15 probands that were solved through likely pathogenic and pathogenic variants in *PRPF31*, only three were included with a suspected autosomal dominant inheritance pattern. Moreover, in proband 067528, a homozygous deletion in the *BBS9* gene was considered to very likely solve the case while in proband 068556, a pathogenic variant in *PRPH2* was considered causative even though pathogenic variants in *PRPF31* were detected in both probands. Especially in cases such as proband 067528, potential syndromic phenotypes should be assessed additionally. These findings underline the added benefit of using this genotype-first approach and thereby considering all prioritized variants, regardless of associated phenotypes or inheritance patterns. Additionally, in 10 cases variants in two or three genes could explain the phenotype of the probands (Supplemental Table 9). For these cases, the phenotype and inheritance pattern provided by the collaborator, as well as the zygosity and the ACMG classification of the variants were taken into consideration. All variants, including the most likely, causative variant, are listed in Supplemental Table 9 and are labeled as “primary” in the verdict column, all additional findings are labeled as “secondary”. In most cases, segregation analysis could already give an indication as to which variants are more likely to solve the proband.

While the smMIPs approach has multiple advantages over other sequencing techniques, such as the ability to sequence large groups of probands in a single sequencing run, the ability to curate and include all desired target regions and the low costs compared to WES or WGS, it does have limitations. For instance, the exact nature of CNVs cannot be identified from the sequencing data, which necessitates additional validation and breakpoint analysis. Moreover, duplications, balanced inversions, and to a lesser extent deletions, are hard to detect since only the genes of interest are sequenced and no comparisons in read counts can be made to neighboring genes on the chromosome. Lastly, while the lower costs compared to WES or WGS, discussed previously in Hitti-Malin *et al*., (2022), makes the smMIPs approach attractive, the extensive infrastructure that is necessary on both sequencing capacity and bioinformatic processing of the data hampers a universal applicability.

In conclusion, the low costs and high-throughput capacity of smMIPs sequencing allowed us to effectively sequence all RP and LCA associated genes and loci in 1,192 probands. Alongside previously published variants, a large group of novel variants could also be detected. As new genetic therapies directed against specific IRD genes are becoming available, easy access to genetic/genomic testing and early genetic diagnosis is of the utmost importance to allow the patient to optimally benefit from these treatments (Black et al., 2021).

## Supporting information

Supplemental figure 1 - RP17 region

Supplemental Table 1 - Gene list including phenotypes

Supplemental Table 2 - DIVs and PEs

Supplemental Table 3 - Positive controls

Supplemental Table 4 - Very likely and possibly solved probands

Supplemental Table 5 - CNVs

Supplemental Table 6 - AR SNVs

Supplemental Table 7 - AD SNVs

Supplemental Table 8 - XL SNVs

Supplemental Table 9 - Probands explained by variants in two or more genes

## Data Availability

All data produced in the present study are available upon reasonable request to the authors

## Acknowledgements

The authors would like to thank all patients involved and their families for taking part in this research. We thank Saskia D. van der Velde-Visser, Ellen A.W. Blokland, Marlie Jacobs-Camps, Anita Roelofs, Michiel Oorsprong, Marcel Nelen, Simon van Reijmersdal, and Martine van Zweeden, and the Radboud Genomics Technology Center for technical assistance. Sarah De Jaegere (Center for Medical Genetics Ghent Belgium) is thanked for her technical support. We also thank Greg Porreca and Eric Boyden at Molecular Loop Biosciences Inc. for their contribution and expertise. This study received financial support from Novartis. We want to acknowledge the Biobank Nodo Hospital Virgen Macarena (Biobanco del Sistema Sanitario Público de Andalucía) integrated in the Spanish National biobanks Network (PT20/00069) supported by ISCIII and FEDER funds for their collaboration in this work. The work in the Chromosomal Genetics Unit is supported by the CHROMOSTEM research platform. This work was supported by grants from Retinitis Pigmentosa Fighting Blindness, Fight for Sight UK (RP Genome Project GR586), Ghent University Special Research Fund (BOF20/GOA/023) (EDB, BPL); EJP RD Solve-RET EJPRD19-234 (EDB, BPL, SB, CR, FC, SR). EDB (1802220N) and BPL (1803816N) are FWO Senior Clinical Investigators of the Research Foundation Flanders (FWO). EDB, BPL, SB, FC, SR are members of ERN-EYE (Framework Partnership Agreement No 739534).

---

Supplemental Figure 1: **RP17 region smMIP distribution**. The entire RP17 region is covered by smMIPs and allows for the detection of duplications that could result in autosomal dominant RP.

Supplemental Table 1: **Genes included in the RP-LCA smMIPs panel**.

Supplemental Table 2: **Known DIVs and PEs included in the RP-LCA smMIPs panel**.

Supplemental Table 3: **Positive control dataset**.

Supplemental Table 4: **Very likely and possibly solved probands**.

Supplemental Table 5: **Copy number variants**.

Supplemental Table 6: **Single nucleotide variants in genes associated with autosomal recessive inheritance**.

Supplemental Table 7: **Single nucleotide variants in genes associated with autosomal dominant inheritance**.

Supplemental Table 8: **Single nucleotide variants in genes associated with X-linked inheritance**.

Supplemental Table 9: **Probands explained by variants in two or more genes**.

