## Supplementary figures and images for "Cost-effective sequence analysis of 113 genes in 1,192 probands with retinitis pigmentosa and Leber congenital amaurosis"

### Supplemental figure 1 - RP17 region

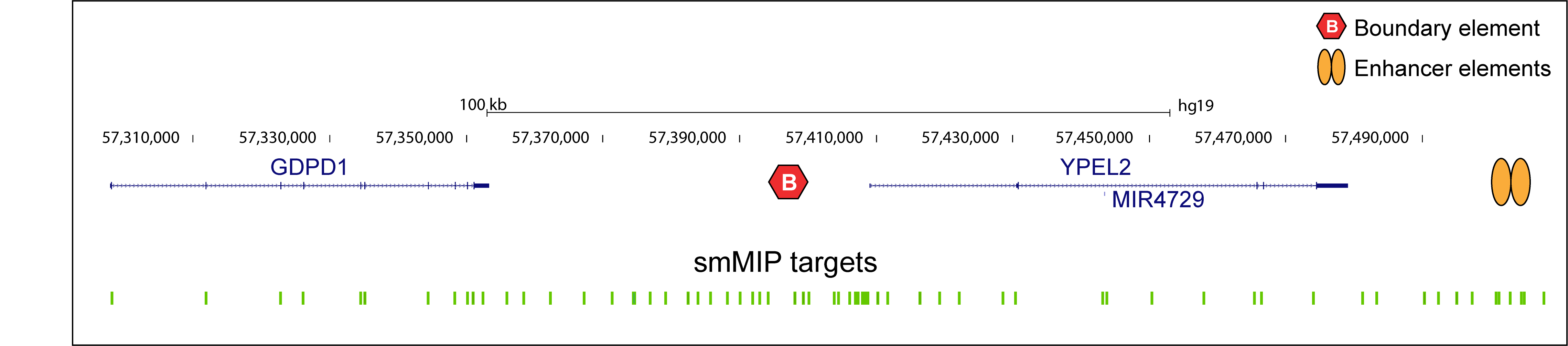
